# Characterization of Entomological Drivers of Malaria Transmission in Five Villages, Keerom Regency, Papua, Indonesia

**DOI:** 10.64898/2025.12.19.25342627

**Authors:** Ismail E Rozi, Lepa Syahrani, Dendi H Permana, Irdayanti, Rusdiyah, Puji B S Asih, Neil F Lobo, Din Syafruddin

## Abstract

**Background:** Keerom Regency remains one of the high-malaria-endemic regencies in Indonesia. Despite the accelerated malaria elimination strategy underway in Papua, malaria transmission in this regency has not declined. To characterize the entomological and human behavioral factors sustaining and driving malaria transmission, a rapid entomological assessment paired with human behavior observations (HBOs) and household surveys was conducted in five villages of Keerom Regency.

**Methods:** Entomological surveys were conducted on three occasions in 2022 and 2023. Human landing catches (HLC), night indoor resting collection, and mosquito larval site surveys were conducted alongside human behavior observations (HBO) and structured household surveys.

**Results:** Six species of *Anopheles* were identified including *Anopheles koliensis*, *An. punctulatus*, *An. hinesorum*, *An. kochi*, *An. bancroftii* and *An. peditaeniatus*. The dominant species were *An. koliensis* and *An. punctulatus*. Outdoor *Anopheles* human biting rates (HBR) averaged 1.4 ± 1.7 bites per person per hour (bph), higher than indoor HBRs of 0.8 ± 1.2 bph. Vector incrimination detected *Plasmodium* DNA in ten *An. koliensis* samples in 2023, including *Plasmodium falciparum* (n=2), *P. vivax* (n=7) and *P. ovale* (n=1). Blood meal analysis showed mixed feeding on humans and dogs, with human blood indices of 58.3% for *An. koliensis* and 66.7% for *An. punctulatus*. HBOs highlighted a substantial gap in indoor protection during the early evening before sleeping. Household surveys identified several drivers of exposure, including the absence of window and door screens, limited indoor residual spraying coverage, low usage of insecticide-treated nets, and unprotected outdoor activity.

**Conclusions:** Members of the *An. punctulatus* group were abundant across all study villages, with *An. koliensis* and *An. punctulatus* being the primary species. Vector incrimination confirmed active malaria transmission. These vectors demonstrated opportunistic feeding behavior, primarily on humans but also on dogs, which may offer an avenue for targeted interventions. Multiple gaps in personal and household protection were identified, both indoors and outdoors. Strengthening indoor protection through increased ITN use, installation of house screens, and evaluation of complementary tools such as spatial repellents may reduce indoor transmission. Community-led larval source management in and around households may help reduce outdoor transmission.

## INTRODUCTION

Indonesia launched its national malaria elimination program in 2009, with the goal of reaching elimination by 2030 [1]. More than 400 of the country’s 514 regencies and municipalities have achieved elimination, yet transmission in Papua Province remains high [2]. Core national strategies such as early diagnosis and prompt treatment, mass distribution of insecticide-treated nets (ITNs), and indoor residual spraying (IRS) [1,3] have not been sufficient to reduce malaria incidence in this region [2]

Because of Papua has distinct epidemiologic and entomologic features [4], an accelerated elimination strategy was introduced in the National Action Plan for 2020–2026. This plan aims to ensure that no regency in Papua remains in the moderate or high transmission category and to reduce the provincial positivity rate to below 5% by the end of 2026 [5]. The strategy includes intensified activities such as mass drug administration (MDA), school-based screening, active case detection by community malaria workers, intermittent preventive treatment in pregnancy (IPTp), and integrated vector control in regencies with annual parasite incidence (API) above 100 per thousand. Supporting activities include monitoring of insecticide resistance, ITNs durability and efficacy, and quality assurance on the therapeutic efficacy study (TES) of anti-malarial drugs in selected regencies.

Keerom Regency has the fifth-highest malaria incidence in Papua Province [6]. Although malaria vector control activities in Keerom Regency, such as mass distribution of ITNs, IRS spraying, and other components of the accelerated program have been implemented, malaria incidence has not declined (Fig. 1). This gap points to a need for entomology-based evaluations. To date, few studies in Papua have incorporated detailed entomological surveillance to assess the effectiveness of vector control implementation.

**Fig. 1.**
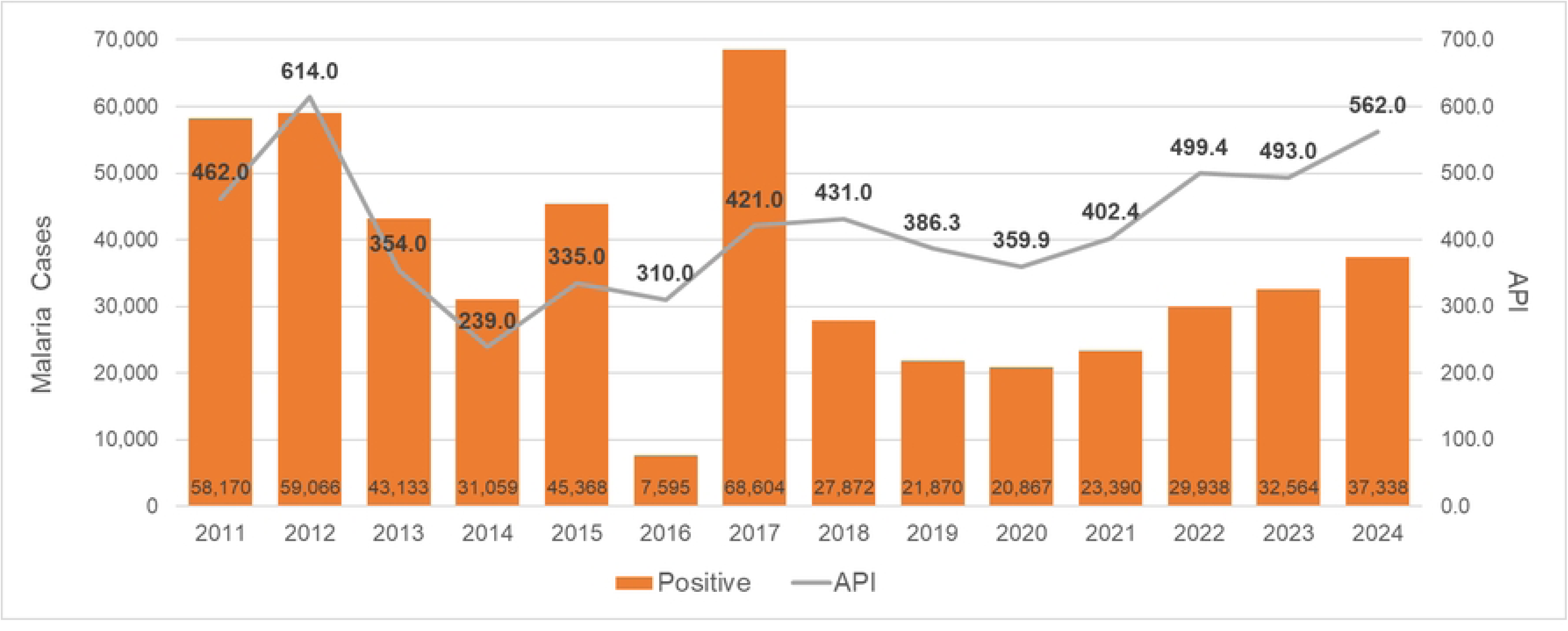
Malaria morbidity rates per year from 2011 to 2024 in Keerom Regency. Source: Keerom Regency Statistics Agency 2014 – 2025.

To address this need, rapid entomological assessments were carried out in eight high-endemic regencies of Papua Province in 2019 and 2021 [7,8]. This study was one of the efforts to identify problems in vector control interventions in the Papua region. These assessments developed a baseline understanding of the transmission system including the gaps in protection towards developing a targeted vector-based intervention strategy. The findings demonstrated that members of the *An. punctulatus* group are the primary vectors, biting both indoors and outdoors. *An. koliensis* and *An. punctulatus* were widespread across ecological settings, whereas *An. farauti* was more common in coastal areas. Larval surveys revealed that many water bodies in and around residential areas supported *Anopheles* breeding. The studies also recommended installing house screening and testing innovative tools, such as spatial repellents, to reduce indoor transmission, as well as community-led larval source management and strengthened social and behavioral change communication to reduce outdoor exposure [7].

While informative, these earlier assessments were limited by short sampling periods and weather constraints [7]. Additional data collected over longer timeframes and at different sites are needed to validate and refine entomological indicators of transmission risk. This study addresses that need by conducting extended entomological surveillance in Keerom Regency. Five villages with high malaria incidence in 2022, as identified through Indonesia’s malaria information system (SISMAL), managed by the ministry of health [9], were selected as study sites. The objective was to characterize local vectors, larval habitats, and human behavior to determine where and when transmission occurs and to identify gaps in current vector control efforts.

## METHODS

### Ethical statement

Ethical review and approval for this study were granted by the Ethics Committee of Research in Health, Medical Faculty of Hasanuddin University, Makassar, Indonesia, Protocols No. 371/UN4.6.4.5.31/PP36/2022 (year 2022), No. 265/UN4.6.4.5.31/PP36/2023 (year 2023), and the extension of No. 311/UN4.6.4.5.31/PP36/2024 (year 2024).

### Site selection

Keerom Regency, has a total population of 74,332, and remains one of the five highest malaria-endemic regencies in Papua Province. In 2024, the regency reported 37,338 malaria cases reported and an API of 562 (Fig. 1) [10]. Keerom lies along the eastern border with Papua New Guinea and consists of 11 subdistricts covering 9,365 km², with more than 60% of the area situated between 400 and 1,500 meters above sea level. Rainfall in 2023 was moderate, with the heaviest rainfall in March and the lowest in November [11].

Field activities were conducted three times during 2022 and 2023, across five villages located in three subdistricts: Sanggaria and Yaturaharja villages (Arso Barat Primary Health Center (PHC)), Ubiyau and Sawanawa villages (Arso Kota PHC), and Pitewi village (Pitewi PHC) (Fig. 2) [12]. The first survey was conducted from September 19^th^ to October 8^th,^ 2022, in four villages (Sanggaria, Yaturaharja, Ubiyau, and Sawanawa). The second survey took place from May 15^th^ to 28^th,^ 2023, in the same four villages (Sanggaria, Yaturaharja, Ubiyau, and Sawanawa), and the third survey was conducted in Pitewi from July 4^th^ to 17^th,^ 2023. Climate data corresponding to the sampling periods were obtained from the nearest meteorological station in Sentani, Jayapura (Fig. 3) [11,13].

**Fig. 2.**
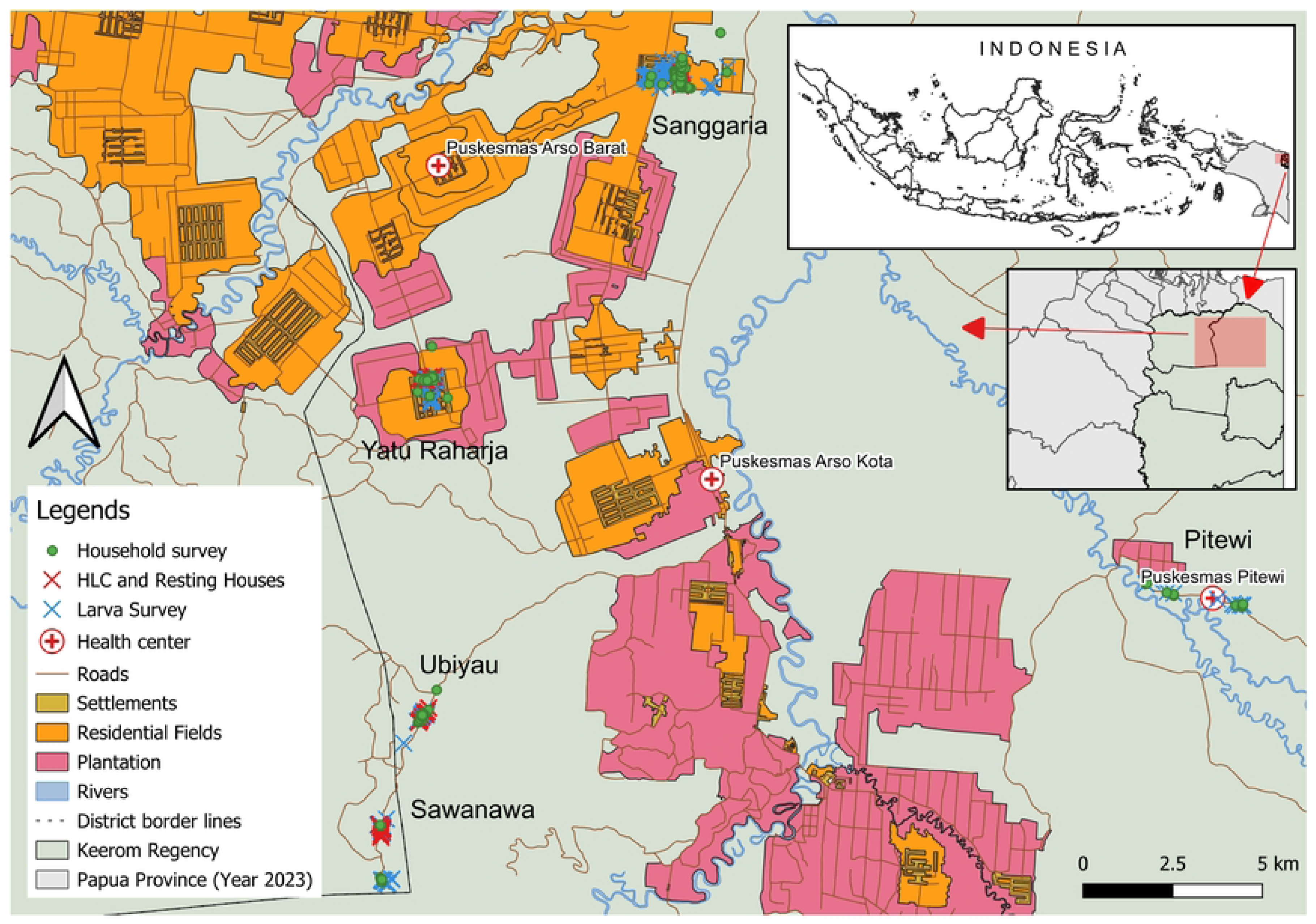
Location of study sites. Map source: Ina Geoportal (https://tanahair.indonesia.go.id/portal-web/) [12].

**Fig. 3.**
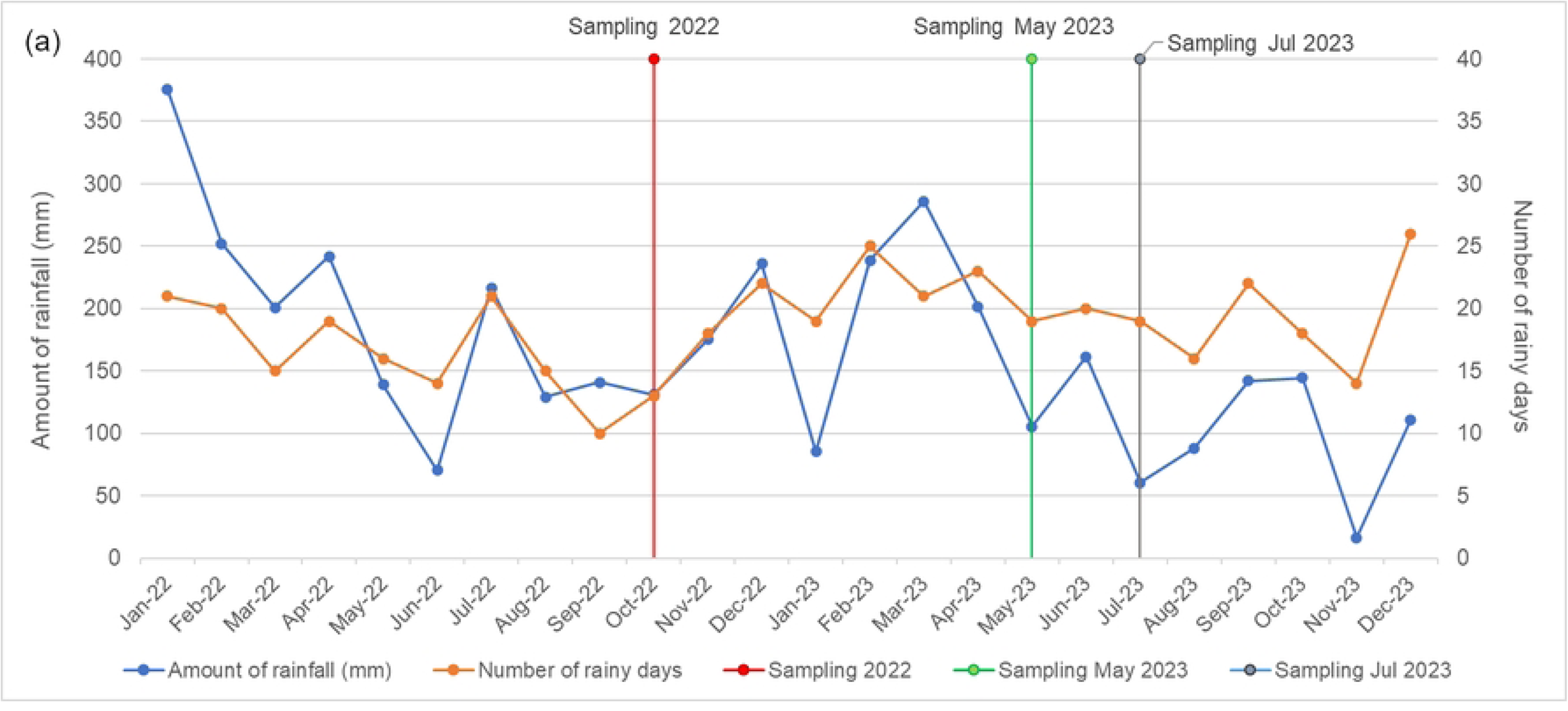
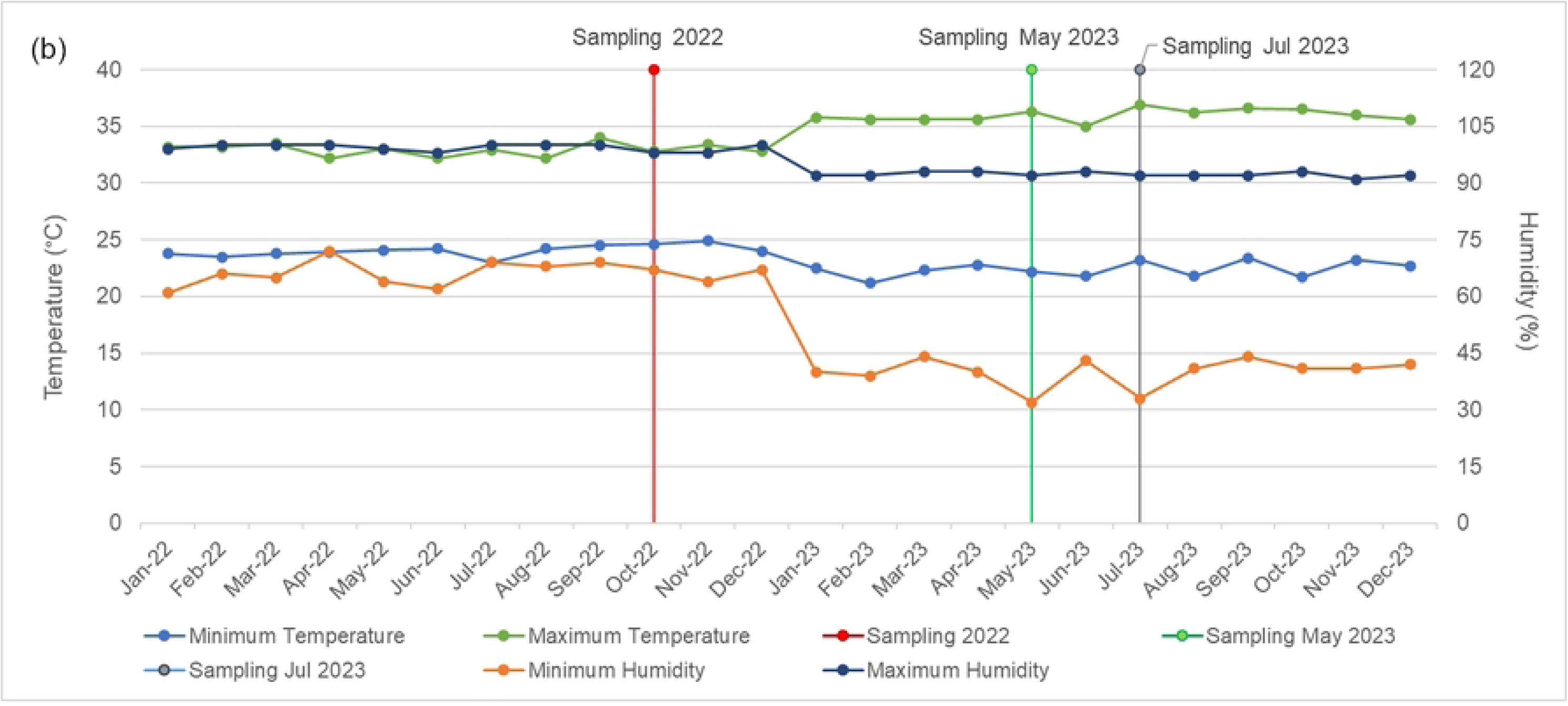
Weather at the time of sampling in Keerom Regency in 2022 and 2023, data of (a) rainfall and rainy days and (b) temperature and humidity. The climate data was taken from the nearest climate observation station (Sentani Climate Observation Station, Jayapura Regency) [11,13].

### Study Population

Study participants were residents living near mosquito sampling sites. Roughly 60% of the Keerom population works in agriculture, forestry, or fisheries. [11]. The villages of Sanggaria and Yaturaharja are largely inhabited by transmigrant communities, while Ubiyau and Sawanawa are primarily populated by Indigenous Papuan residents. Pitewi has a mix of local and non-local populations. [14–16]. Cultural practices and nighttime behaviors vary across these communities, influencing exposure to mosquito bites..

### Entomological endpoints

Measured entomological endpoints included the host-seeking behavior (indoors and outdoors), human biting rate (HBR, bites per person per night), indoor resting density (IRD), sporozoite rate, and the entomological inoculation rate (EIR) (infectious bites per person per night). The sporozoite rate was calculated as the number of mosquitoes infected with *Plasmodium* divided by the number of mosquitoes examined. EIR was calculated as the product of HBR and the sporozoite rate. Adult mosquitos were sampled using human landing catch (HLC) and indoor resting collections. Adult vector densities were assessed using standardized human-landing catches (HLC) and IRDs. Immature stages were sampled daily across potential larval habitats. Adult sampling supported spatial and temporal analyses of species abundance and diversity. [17,18].

### Mosquito collections

#### Human landing catch (HLC)

HLCs were conducted from 18:00 to 06:00 in four sentinel houses per night. Each site was sampled for two nights, except Ubiyau and Sawanawa in 2022, where only one night of sampling was possible. Eight trained adult volunteers (four indoors, four outdoors) participated after providing written informed consent. Human landing rates (HLR) were used as proxies for HBR and were calculated as bites per person per hour or per night.

#### Indoor resting collections

Indoor resting mosquitoes were sampled using manual aspirators every two hours from 18:00 to 06:00 in four houses per night. Sampling occurred on the same nights as HLCs but in different houses. Each site was sampled on two nights, except Ubiyau and Sawanawa in 2022, where indoor resting collections were not performed. IRD was calculated as the number of *Anopheles* per house per night.

#### Mosquito larval collection

Larval surveys targeted water bodies adjacent to human dwellings and surrounding the village. All potential breeding sites were surveyed and classified into six habitat types: ponds, ditches/gutters, seepage/springs/wells, rain pools or puddles, stream margins, and swamps. Habitat positivity was recorded as the presence of Anopheles larvae. Larval density was calculated as total larvae per total dips. Habitat characteristics examined included type, sun exposure, water stability, flow rate, and vegetation [19,20].

### Blood meal analysis

Blood-fed Anopheles collected through indoor resting methods were randomly selected for blood meal source identification. PCR was performed using primers specific for human, dog, pig, goat, and cattle DNA [21]. PCR products were visualized by agarose gel electrophoresis. Blood meal sources were determined from amplicon length. The human blood index (HBI) represented the proportion of blood meals derived from humans [22].

### Species identification

All adult Anopheles specimens were identified morphologically using the J. Bonne-Wepster et al. (1953) [23] key. Species in the *Punctulatus* complex were differentiated using the Rozeboom and Knight (1946) [24] and Bryan (1974) [25] morphological keys. A subset of adult specimens from all species and sites was confirmed molecularly using ITS2 sequencing [26,27]. Larvae were also identified molecularly. Sequences were compared with reference sequences using BLASTn [28]. Agreement between morphological and molecular identification was evaluated using sensitivity and positive predictive value metrics.

### Human behavior observations (HBO)

HBOs were conducted hourly from 18:00 to 06:00 in the same houses used for HLCs. Observations captured human movement, location (indoor or peri-domestic), and ITN use. Data were collected by HLC volunteers using paper forms and covered the immediate area around each structure. Volunteers themselves were excluded from HBO analyses. Human exposure was calculated by combining human presence, ITN use, and vector biting patterns [29–32].

### Household survey and identification of gaps in protection

Household surveys were conducted with support from trained health workers using the ODK Collect mobile application [33]. Each village aimed to enroll 20–30 respondents aged 15 years or older. The survey captured housing characteristics, socioeconomic indicators, nighttime behavior, mosquito protection practices, and recent malaria history [29,34]. Gaps in protection [35] were assessed across several domains: exposure-adjusted biting rates, house structure and screening, ITN distribution and use, IRS coverage, and nighttime behaviors. Gaps were categorized as minor (+), moderate (++), or major (+++). Associations with village and socioeconomic status were tested using Pearson’s chi-square or Fisher’s exact tests.

### Data Analysis

Data from surveys and larval collections were captured using ODK and uploaded to a secure server [33,36]. Quantitative analyses, including descriptive statistics, figures, and statistical tests, were performed in Microsoft Excel and RStudio (R version 4.3.3) [37,38]. Socioeconomic status was estimated using a wealth index derived from principal component analysis of 22 asset variables. Households were ranked into quintiles for comparison with other variables [8].

## RESULTS

### Adult *Anopheles* collection results

Six genera of mosquitoes were identified morphologically and included samples from *Anopheles*, *Aedes*, *Culex*, *Armigeres*, *Mansonia,* and *Coquillettidia*. A total of 5450 adult mosquitoes were collected, consisting of 3,797 mosquitoes from HLC and 1,653 from indoor resting collections. Of the *Anopheles* samples (n = 2,099), 1,704 were obtained from HLCs and 395 from indoor resting collections. Morphological identification revealed that most *Anopheles* samples belonged to the *Punctulatus* Group (94.8%); and included *Anopheles koliensis* (74.8%), *An. punctulatus* (19.0%) and *An. farauti* complex (1.0%). There was one sample of *An. bancroftii* from a HLC, and only one *An. kochi* specimen obtained resting indoors. Indoor resting collections resulted in 395 *Anopheles* samples. However, in 2022, only one *Anopheles* sample was. collected (*Anopheles koliensis*) (Table 1). Molecular species identification of the named *An. bancroftii* and *An. kochi* samples was based on *ITS2* barcoding that identified the closest sequences (date of alignment: 2 May 2025) were from *An. bancroftii* with GenBank ID: MT740912.1 (identity 87.82% of 100% coverage) and *An. kochi* OR290055.1 (identity 99.79% of 100% coverage) respectively.

**Table 1.** Morphologically identified total number of adult *Anopheles* species per village.

| Collection<br>Method | Species | Villages |  |  |  |  |  |  |  |  |  | Total |  |
| --- | --- | --- | --- | --- | --- | --- | --- | --- | --- | --- | --- | --- | --- |
|  |  | Sanggaria |  | Yaturaharja |  | Ubiyau |  | Sawanawa |  | Pitewi |  |  |  |
|  |  | 2022 | 2023 | 2022 | 2023 | 2022 | 2023 | 2022 | 2023 | 2022 | 2023 | 2022 | 2023 |
| HLC | *An. koliensis* | 45 | 540 | 16 | 152 | 30 | 258 | 2 | 55 | - | 163 | 93 | 1168 |
|  | *An. punctulatus* | 40 | 4 | 5 | 0 | 43 | 3 | 105 | 7 | - | 118 | 193 | 132 |
|  | *An. hinesorum* | 0 | 4 | 1 | 1 | 2 | 6 | 0 | 3 | - | 2 | 3 | 16 |
|  | *An. kochi* | 0 | 0 | 0 | 0 | 0 | 0 | 0 | 0 | - | 0 | 0 | 0 |
|  | *An. bancroftii* | 0 | 0 | 0 | 0 | 0 | 0 | 0 | 1 | - | 0 | 0 | 1 |
|  | Undetermined | 26 | 0 | 21 | 0 | 18 | 0 | 18 | 0 | - | 15 | 83 | 15 |
| Night<br>Indoor<br>Resting | *An. koliensis* | 0 | 37 | 1 | 19 | - | 146 | - | 7 | - | 86 | 1 | 295 |
|  | *An. punctulatus* | 0 | 0 | 0 | 0 | - | 3 | - | 1 | - | 75 | 0 | 79 |
|  | *An. hinesorum* | 0 | 0 | 0 | 0 | - | 0 | - | 2 | - | 0 | 0 | 2 |
|  | *An. kochi* | 0 | 0 | 0 | 0 | - | 0 | - | 1 | - | 0 | 0 | 1 |
|  | *An. bancroftii* | 0 | 0 | 0 | 0 | - | 0 | - | 0 | - | 0 | 0 | 0 |
|  | Undetermined | 4 | 0 | 2 | 3 | - | 0 | - | 1 | - | 7 | 6 | 11 |
| Total |  | 115 | 585 | 46 | 175 | 93 | 416 | 125 | 78 | - | 466 | 379 | 1720 |

Of the 45 randomly selected *Anopheles* that were examined molecularly, the sensitivity of morphological identification for *An. koliensis* was only 35.9%, although the positive predictive value was perfect at 100%. Most of the mismatches were mosquitoes that had been called *An. punctulatus* or members of the *An. farauti* complex, which were later identified as *An. hinesorum*. In contrast, both *An. punctulatus* and *An. hinesorum* showed 100% sensitivity, but their PPVs were low at 28.6% and 11.8%.

A total of 107 *Anopheles* (5.1%) could not be identified morphologically because they were damaged, moldy, or missing parts. Twenty-eight of these were included in molecular testing. From these, we identified 19 *An. koliensis* and nine *An. punctulatus*.

The hourly pattern of biting (Fig. 4) shows that *Anopheles* were active from the start of HLC at 18:00 until 06:00, with a clear peak between midnight and 01:00. The mean hourly HBR was 1.1 ± 1.2 bites per hour (range 0.0 to 5.5). Outdoor biting pressure was higher than indoor, at 1.4 ± 1.7 bph (0.0 to 8.8) compared to 0.8 ± 1.2 bph (0.0 to 6.8). A Welch two-sample t-test confirmed the difference between indoor and outdoor biting (p = 0.0001).

**Fig. 4.**
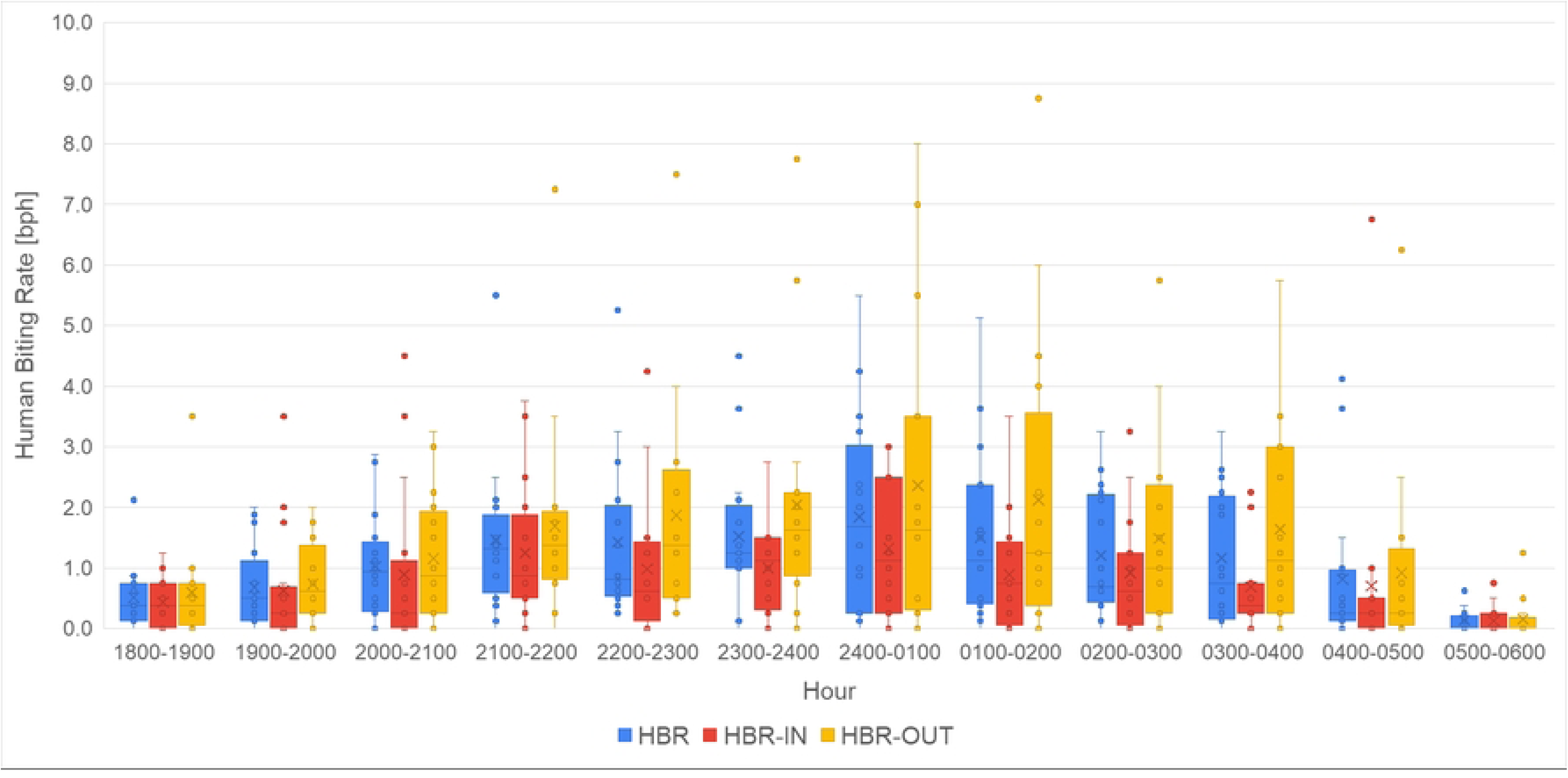
*Anopheles* biting behaviors based on HLC collections in five villages.

Figure 5 shows the distribution of HBR and indoor resting density (IRD) across sites and years. Except for Sawanawa, all 2023 HBRs were higher than in 2022. The mean HBR in 2022 was 7.8 ± 8.1 bites per night (0.0 to 26.5), compared to 16.7 ± 16.2 in 2023 (0.5 to 71.0). A one-way ANOVA found significant differences across sites and years (p = 2.8 x 10^-5). Tukey HSD highlighted strong differences for Sanggaria 2023 (SG23) compared to YT22, SW23, and SG22. Mean IRD across all sites was 7.1 ± 13.2 mosquitoes per house per night (0 to 81). Pitewi and Ubiyau reported the highest IRDs. IRD and overall HBR were not correlated (Pearson p = 0.3618).

**Fig. 5.**
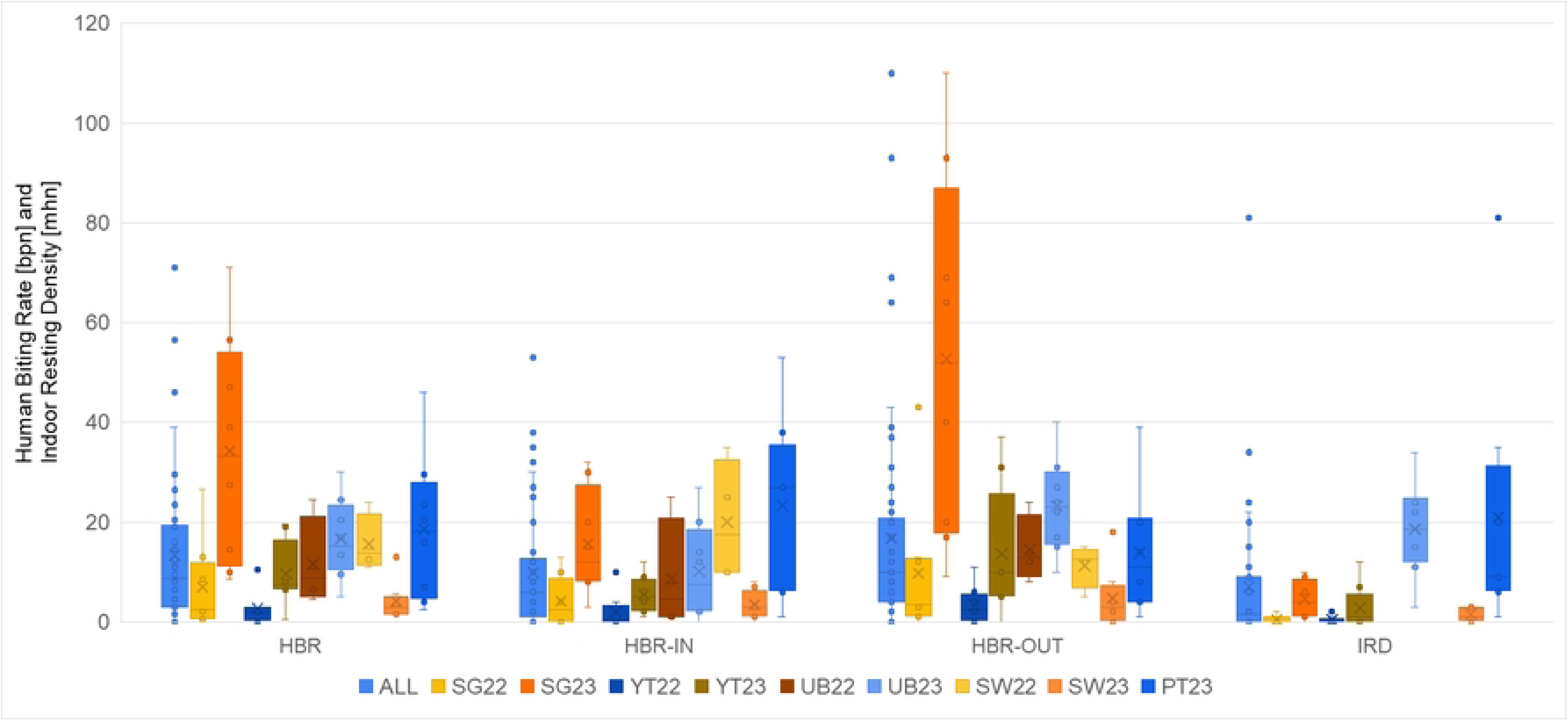
Human biting rate (HBR) and Indoor resting densities (IRD) per sentinel house per night from each five study sites in year 2022 and 2023. SG22 is mosquito sampling in Sanggaria year 2022, SG23: Sanggaria year 2023, YT22: Yaturaharja year 2022, YT23: Yaturaharja year 2023, UB22: Ubiyau year 2022, UB23: Ubiyau year 2023, SW22: Sawanawa year 2022, SW23: Sawanawa year 2023, and PT23: Pitewi year 2023.

Screening for *Plasmodium* DNA in salivary glands from five villages (2022–2023) showed that out of 1,933 adult *Anopheles*, ten samples from 2023 were positive. These included two *P. falciparum*, seven *P. vivax*, and one *P. ovale*. Six positives were from HLC (including all *P. falciparum* and *P. ovale*, plus three *P. vivax* from Sanggaria and Pitewi) (Table 2). Four *P. vivax* positives from Ubiyau were from indoor resting collections. All infected mosquitoes were *An. koliensis*. In 2023, Sanggaria had the highest HBR (34.3 bpn), while Ubiyau had the highest sporozoite rate (1.5%) and the highest EIR (0.25 infectious bites per person per night, or an annual EIR of 90).

**Table 2.** Plasmodium DNA detection in *Anopheles* collected from five villages.

| Village | Year | *Anopheles* species | Collection Method | Positive plasmodial DNA |  |  | Uninfected | Sporozoite Rate | HBR*<sup>4</sup> (bpn) | EIR*<sup>5</sup> (ibpn) | Annual EIR |
| --- | --- | --- | --- | --- | --- | --- | --- | --- | --- | --- | --- |
|  |  |  |  | Pf*<sup>1</sup> | Pv*<sup>2</sup> | Po*<sup>3</sup> |  |  |  |  |  |
| Sanggaria | 2022 | - | - | 0 | 0 | 0 | 88 | 0.0% | 6.9 | 0.00 | 0.0 |
| Yaturaharja | 2022 | - | - | 0 | 0 | 0 | 22 | 0.0% | 2.7 | 0.00 | 0.0 |
| Ubiyau | 2022 | - | - | 0 | 0 | 0 | 76 | 0.0% | 11.6 | 0.00 | 0.0 |
| Sawanawa | 2022 | - | - | 0 | 0 | 0 | 107 | 0.0% | 15.6 | 0.00 | 0.0 |
| Sanggaria | 2023 | *An. koliensis* | HLC-outdoor | 0 | 2 | 1 | 563 | 0.5% | 34.3 | 0.18 | 66.3 |
| Yaturaharja | 2023 | - | - | 0 | 0 | 0 | 166 | 0.0% | 9.6 | 0.00 | 0.0 |
| Ubiyau | 2023 | *An. koliensis* | HLC-indoor,<br>HLC-outdoor | 2 | 0 | 0 | 400 | 1.5% | 16.7 | 0.25 | 90.0 |
|  |  |  | Night Resting | 0 | 4 | 0 |  |  |  |  |  |
| Sawanawa | 2023 | - | - | 0 | 0 | 0 | 73 | 0.0% | 4.1 | 0.00 | 0.0 |
| Pitewi | 2023 | *An. koliensis* | HLC-outdoor | 0 | 1 | 0 | 429 | 0.2% | 18.6 | 0.04 | 15.8 |

Notes:
\*1 *Plasmodium falciparum*
\*2 *Plasmodium vivax*
\*3 *Plasmodium ovale*
\*4 Human biting rate (bpn: bites per person per night)
\*5 Entomological inoculation rate (ibpn: infectious bites per person per night)

We identified blood meal sources from 47 abdominal samples collected from indoor resting mosquitoes. Among the five blood food sources examined—human, dog, pig, goat, and cow— blood meals were only found from humans and dogs. Electrophoresis images showed blood meals solely from dogs (680 bp) in 15 samples and humans (334 bp) in 23 samples. There were also six samples with mixed blood meals from humans and dogs, while three samples did not yield any DNA amplification. Table 3 summarizes these findings. Except for two mosquitoes that could not be identified to species, all were members of the Punctulatus group (*An. koliensis* or *An. punctulatus*). The human blood indices were 58.3% for *An. koliensis* and 66.7% for *An. punctulatus*.

**Table 3.** Results of blood meals analysis of totally 47 blood samples from five villages.

| Village | Blood Meals |  |  |  | *Anopheles* species |  |  | Total Samples |
| --- | --- | --- | --- | --- | --- | --- | --- | --- |
|  | Human | Human and Dog | Dog | Unknown | *An. koliensis* | *An. punctulatus* | *Anopheles* sp. |  |
| Sanggaria | 4 | 0 | 0 | 0 | 2 | 0 | 2 | 4 |
| Yaturaharja | 7 | 0 | 0 | 1 | 8 | 0 | 0 | 8 |
| Ubiyau | 6 | 1 | 6 | 0 | 12 | 1 | 0 | 13 |
| Sawanawa | 1 | 1 | 2 | 0 | 3 | 1 | 0 | 4 |
| Pitewi | 5 | 4 | 7 | 2 | 11 | 7 | 0 | 18 |
| Total | 23 | 6 | 15 | 3 | 36 | 9 | 2 | 47 |

### Larval surveys

Approximately 181 water bodies were surveyed in residential areas and nearby surroundings and grouped into six habitat types (Table 4). Habitat positivity for *Anopheles* larvae is presented by sampling year (2022 and 2023) (Fig. 6). The most common habitat types were ditches or gutters, puddles or rain pools, and ponds. Puddles and rain pools formed in animal footprints, vehicle ruts, and various discarded containers such as tires, buckets, and drums. A swamp habitat was found only in Sanggaria and was better described as a marsh. *Anopheles* habitat indices varied across habitat types, even within the same village or sampling period. Overall, habitat indices were higher in 2023 than in 2022, except in Sanggaria. For ponds specifically, suitability for Anopheles did not change much between years in Ubiyau and Sawanawa. In Sanggaria, Yaturaharja, and Pitewi, ponds, ditches or gutters, and puddles or rain pools all served as notable larval habitats.

**Fig. 6.**
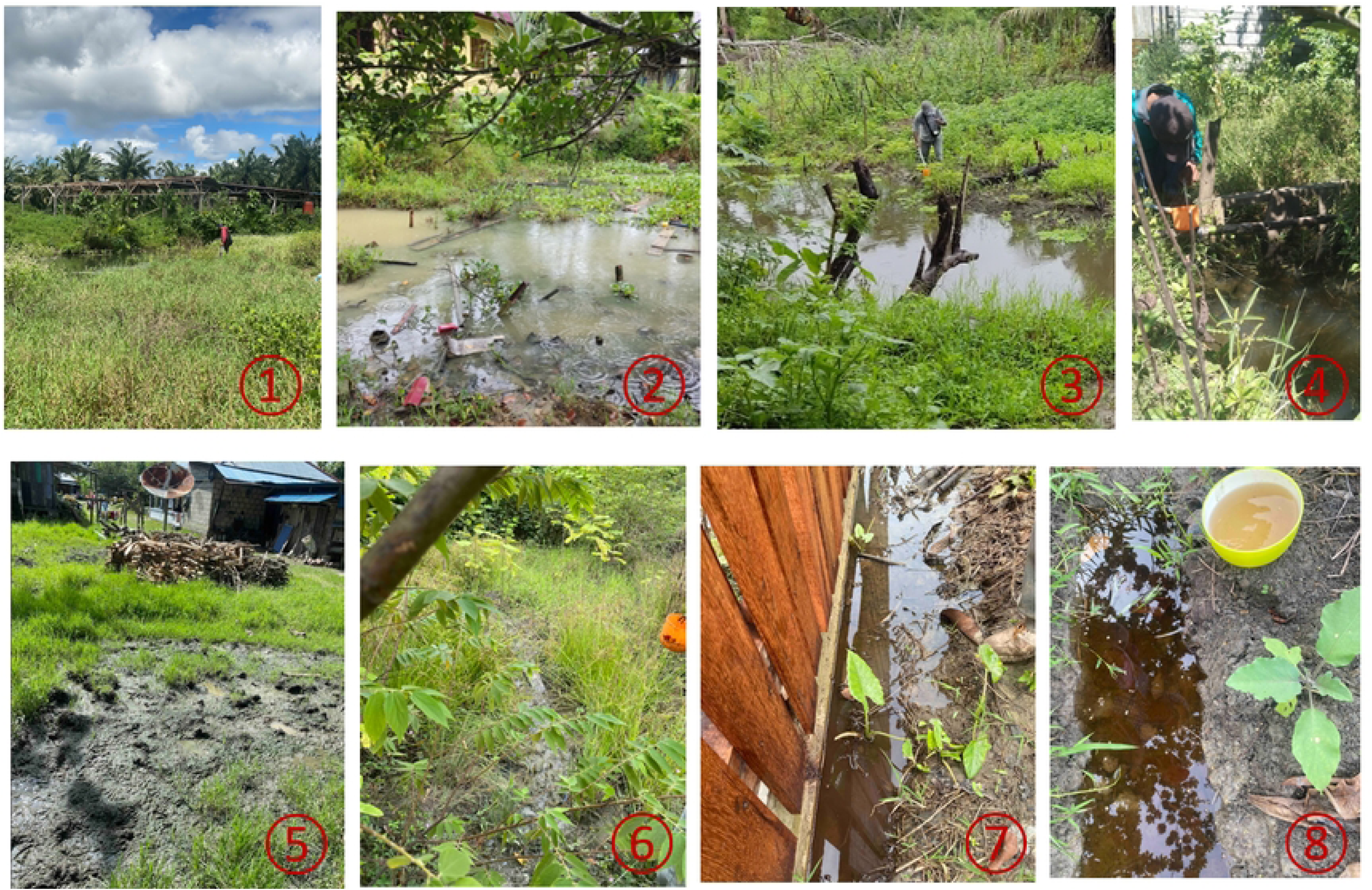
*Anopheles* larval habitat types, ① was a marsh, ② and ③ ponds, ④ ditch/gutter, ⑤ cow hoof prints, and ⑥ – ⑧ were puddles/rain pools.

**Table 4.** The habitat index of *Anopheles* mosquito larvae according to habitat type (2022 and 2023 sampling).

| Habitat type | Sanggaria |  | Yaturaharja |  | Sawanawa |  | Ubiyau |  | Pitewi |  | Total |  |
| --- | --- | --- | --- | --- | --- | --- | --- | --- | --- | --- | --- | --- |
|  | 2022 | 2023 | 2022 | 2023 | 2022 | 2023 | 2022 | 2023 | 2022 | 2023 | 2022 | 2023 |
| ponds | 33.3% (6) | 0.0% (4) | 0.0% (4) | - | 20.0% (5) | 33.3% (9) | 66.7% (3) | 66.7% (3) | - | 0.0% (3) | 27.8% (18) | 26.3% (19) |
| ditches/gutters | 7.7% (26) | 0.0% (14) | 0.0% (14) | 50.0% (2) | 75.0% (4) | 33.3% (3) | 0.0% (2) | 0.0% (2) | - | 42.9% (7) | 10.9% (46) | 17.9% (28) |
| spring/wells | 0.0% (5) | 0.0% (1) | - | - | - | - | 0.0% (3) | - | - | - | 0.0% (8) | 0.0% (1) |
| puddles/rain pools | 0.0% (10) | 16.7% (6) | 0.0% (8) | 66.7% (3) | 23.1% (13) | 36.4% (11) | 25.0% (4) | - | - | 50.0% (2) | 11.4% (35) | 36.4% (22) |
| stream margin | - | - | - | - | 0.0% (1) | - | - | - | - | - | 0.0% (1) | - |
| swamps | 50.0% (2) | 100.0% (1) | - | - | - | - | - | - | - | - | 50.0% (2) | 100.0% (1) |
| Total | 10.2% (49) | 7.7% (26) | 0.0% (26) | 60.0% (5) | 30.4% (23) | 34.8% (23) | 25.0% (12) | 40.0% (5) | - | 33.3% (12) | 13.6% (110) | 26.8% (71) |

Note: The number in brackets is the total amount of water bodies observed.

Examples of habitat types with a positive index for *Anopheles* larvae are in Fig. 6. Roughly 94.1% of water bodies positive for Anopheles larvae were located near settlements. Larval density analysis using the Kruskal-Wallis Test showed no significant differences across the six habitat types (p = 0.07194). Tests on habitat conditions showed no significant effects for exposure (p = 0.4554), stability (p = 0.691), or water flow (p = 0.9595). Vegetation was the only factor associated with differences in larval density (p = 0.03209).

Molecular species identification was not conducted for all larval sites. Only larvae from a single site, a marsh in Sanggaria where collections were abundant, were analyzed. Of the total 24 larval samples, 13 samples were identified as *An. peditaeniatus* and 11 samples as *An. koliensis*.

### Human behavior observation

Table 5 summarizes exposure to *Anopheles* bites across the five study villages in 2022 and 2023. HBO data from Pitewi were excluded because of missing values. In most cases, outdoor biting exceeded indoor biting, except in Sawanawa in 2022 and Pitewi in 2023. Even so, average indoor HBRs were still notable and should not be considered low. Across villages, only 52.2% of residents used ITNs during peak host-seeking hours (range 34.6 to 65.5%). After adjusting for human behavior, 45.9% of the population (range 24.4 to 62.2%) were considered protected (by ITNs) from *Anopheles* bites. Behavior-adjusted exposure for unprotected individuals was higher indoors than outdoors, at 33.5% and 20.6% respectively. Figure 7 shows human behavior patterns alongside indoor and outdoor biting rates, and the resulting adjusted biting rates for three villages (Sanggaria, Ubiyau, and Sawanawa) in 2022 and 2023. HBRs were higher in 2023 than in 2022 in every village except Sawanawa. As a result, the bite risk for unprotected individuals increased in 2023 (Table 5). In Sanggaria, ITN use increased from 35% in 2022 to 56% in 2023, but the risk of bites before people actually went to sleep remained higher in 2023 (Fig. 7a). In Ubiyau, behavior-adjusted bite risk for an unprotected individual decreased in 2023 (Fig. 7b). Figure 7c shows the situation in Sawanawa, where higher ITN use and a lower HBR in 2023 reduced bite risk for unprotected individuals. Across settings, there were consistent gaps in protection before people go to sleep under ITNs (Figure 7a–c).

**Fig. 7.**
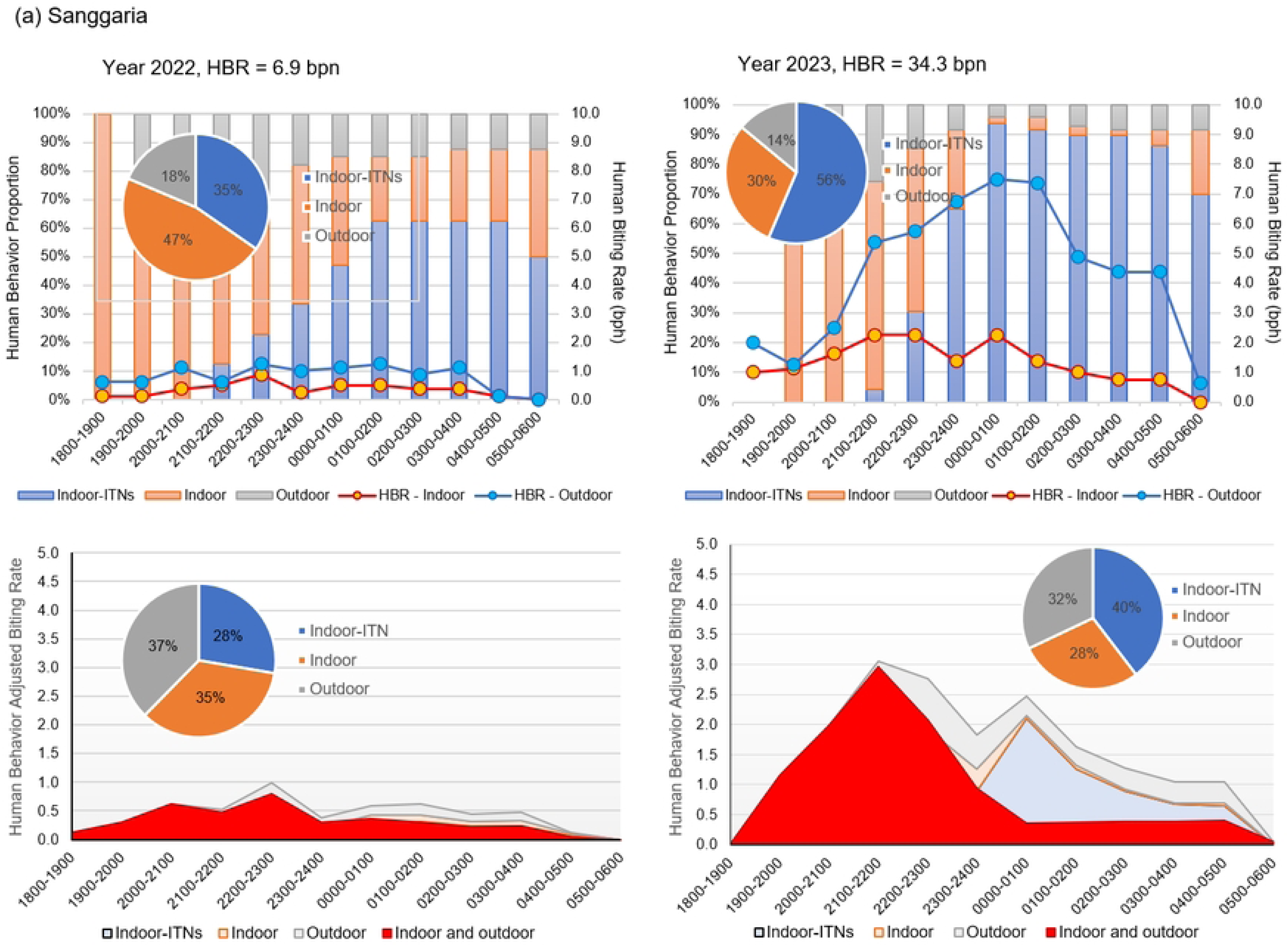
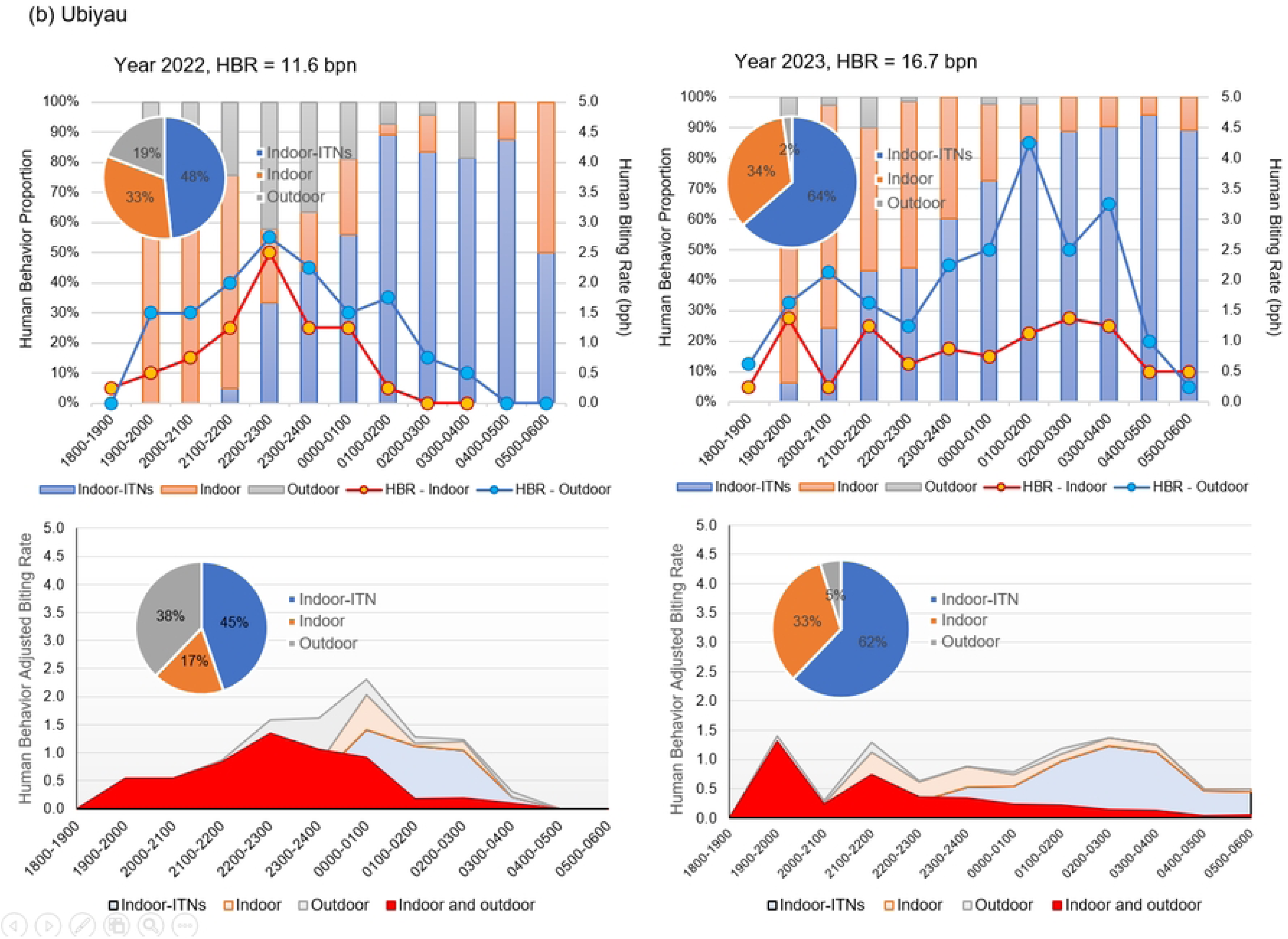
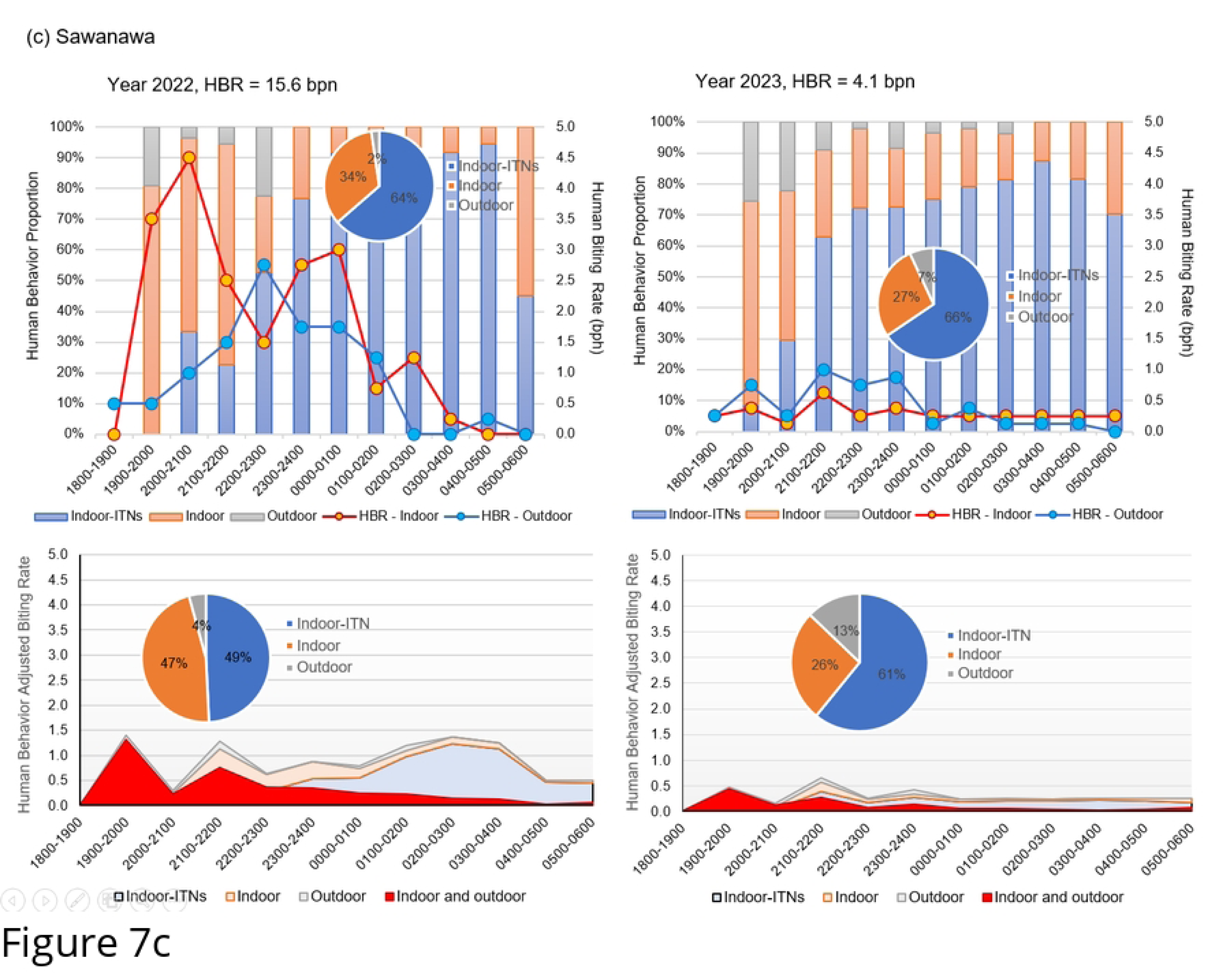
The human behavior adjusted biting rate for unprotected individual from (a) Sanggaria, (b) Ubiyau, and (c) Sawanawa in year 2022 and 2023.

**Table 5.** the exposure of *Anopheles* bites from 5 villages, in 2022 and 2023.

| Biting Behavior | Sanggaria<br>(2022) | Yaturaharja<br>(2022) | Ubiyau<br>(2022) | Sawanawa<br>(2022) | Sanggaria<br>(2023) | Yaturaharja<br>(2023) | Ubiyau<br>(2023) | Sawanawa<br>(2023) | Pitewi<br>(2023) | All Villages |
| --- | --- | --- | --- | --- | --- | --- | --- | --- | --- | --- |
| Directly Measured Biting |  |  |  |  |  |  |  |  |  |  |
| HBR (bpn) | 6.9 | 2.7 | 11.6 | 15.6 | 34.3 | 9.6 | 16.7 | 4.1 | 18.6 | 13.3 |
| Indoor HBR (bpn) | 4.1 | 2.0 | 8.8 | 20.0 | 15.8 | 5.5 | 10.1 | 3.5 | 23.4 | 9.8 |
| Outdoor HBR (bpn) | 9.8 | 3.4 | 14.5 | 11.3 | 52.8 | 13.6 | 23.3 | 4.8 | 13.9 | 16.8 |
| Proportion biting indoors | 29.7% | 37.2% | 37.6% | 64.0% | 23.0% | 28.8% | 30.3% | 42.4% | 63.0% | 38.8% |
| Behavior- Adjusted Exposure - ITN-User |  |  |  |  |  |  |  |  |  |  |
| Proportion of ITN-user | 34.6% | 39.5% | 61.2% | 48.2% | 56.4% | 50.1% | 65.5% | 63.6% | - | 52.2% |
| Proportion of all vector bites prevented by using an ITN | 27.7% | 24.4% | 49.2% | 44.7% | 39.6% | 42.1% | 60.9% | 62.2% | - | 45.9% |
| Behavior-Adjusted Exposure for unprotected individual |  |  |  |  |  |  |  |  |  |  |
| Proportion of vector bites occurring outdoor | 37.6% | 7.1% | 4.3% | 37.8% | 32.0% | 29.7% | 12.9% | 4.9% | - | 20.6% |
| Proportion of vector bites occurring Indoor | 34.7% | 68.5% | 46.5% | 17.5% | 28.3% | 28.2% | 26.3% | 32.9% | - | 33.5% |
| Proportion of vector bites occurring indoor and outdoor | 72.3% | 75.6% | 50.8% | 55.3% | 60.4% | 57.9% | 39.1% | 37.8% | - | 54.1% |

### Household survey

Totally 201 households from five villages were involved into household survey in this study, consisted of Sanggaria = 76, Yaturaharja = 49, Sawanawa = 17, Ubiyau = 29 and Pitewi = 30 (Table S1-1 in the Supplementary File 1). The interviewees were mostly heads of the household or their spouses, with a female-to-male ratio of 1.5:1, ranging from 15 to 78 years old (average 41.8 ± 13.4 years). Their last educations were mostly from primary/junior high school, 42.8% and senior high school or above, 43.8%. The household’s primary occupation as a farmer was higher in all five villages (average 37.3%); however, in Sawanawa, 70.6% of them were farmers. Other occupations were merchant, laborer-technician, woodcutter, and civil servant. Table S1-1.B shows that one household had an average of approximately four members, with fewer than one child under five years old. The average number of de facto household members the previous night was 3.8, consistent across all five villages. Wealth index analysis shows that Sanggaria and Yaturaharja had higher proportions of wealthier households (4th and 5th quantiles), while Ubiyau and Sawanawa had more poorer households (1st and 2nd quantiles). The structure of houses in the five villages was 70% built from cement bricks or concrete, and 29% were wooden houses. Houses with wooden floors accounted for 19.4% of the surveyed households, with two-thirds being stilt houses. About 80% of houses had ceramic or cement floors, and one-third of these were raised above the ground. Most houses had two doors and more than five windows, but only a few had mosquito screens installed (except for houses in Pitewi). Additionally, of the 93.5% of houses with eaves, only 17.0% were partly screened, and 18.1% had screens to prevent mosquitoes from entering. Nearly all houses had electricity supplied by the State Electricity Company.

Table S1-1.C shows the existence of malaria cases that had occurred previously. The malaria cases that had occurred in Sanggaria were the lowest cases, but there were still cases occurring in the last month or even the previous 2 weeks before the interview. Meanwhile, most malaria cases occurred in Ubiyau, with 48% of the recent cases happening in the last month. In Sawanawa, 64.7% of households suffered from malaria, although 63.6% of the recent cases were from the last month, but none since the previous two weeks. Most of the interviewees reported that they learned about malaria from their physician’s examination results, microscopy, RDT results, or symptoms. Only a few malaria-related deaths were reported from Sanggaria in the past two years.

Result of the efforts against mosquito bites is shown in Table S1-1.D. The average IRS coverage is 36.8% of the total surveyed houses, but it was not completely or continuously sprayed in all villages. Ubiyau had the highest IRS coverage at 86.2%, with two-thirds of the houses sprayed less than three months ago, while in Sawanawa, IRS spraying was never done at all. Mostly in all villages IRS was sprayed by health workers, but in Ubiyau it sprayed by NGO (non-government organization). The houses mostly sprayed in Sanggaria, Yaturaharja, and Pitewi occurred more than six months ago.

The average ITN-access in these five villages was 86.1%, indicating high ITN coverage. The average number of bedrooms per house was similar to the number of ITNs. Except in Pitewi, the use of ITNs for everyone in the household was common in all villages. Likewise, most inhabitants reported sleeping under ITNs last night. The ITNs distributed were received less than a year ago and are still in good condition. The proportion of mosquito repellent use in the house was also quite high (average 67.2%), with Sawanawa having the lowest usage at 23.5%. Mosquito coils and sprays were the most common indoors, while the use of personal mosquito repellents was very low.

Table S1-1.E shows the survey results for residents’ nighttime activities. Dinner was mostly done before 7:00 PM in Sawanawa, but in the other villages, the evening meal was commonly consumed between 7:00 PM and 9:00 PM. Evening meals and post meal resting were mostly conducted indoors. Approximately one-third of the interviewees reported that they usually went outside at night, with double the numbers reportrd for Sawanawa and Pitewi. Most of them went out to the neighborhoods, where some of them went hunting or fishing. Moreover, an average of 70.3% of interviewees were staying outdoors for more than one hour, and, except for Sanggaria, more than two-thirds of them were also not using mosquito repellent outdoors. Sleeping behaviors show that the majority of inhabitants slept after 21.00 and woke up before 06.00, and most of them used ITNs.

The knowledge, attitude, and practices (KAP) result is presented in Table S1-1.F. Almost all the interviewees had known about malaria, where the information mainly was received from the local health center or health workers, and also from their neighborhoods. Moreover, they also knew that malaria was transmitted by a mosquito that had already been infected with malaria. The interviewees commonly reported experiencing at least four symptoms of malaria, including shaking chills, fever, nausea, dizziness, and vomiting, with more than half mentioning these symptoms. The most frequently cited prevention methods in the five study villages were sleeping under mosquito nets and burning mosquito coils (spatial repellants). Other common prevention strategies included wearing long-sleeve shirts and trousers and draining stagnant water, though these were not mentioned in Sawanawa. These prevention methods were chosen mainly because they are low-cost, easy to use, and effective at preventing mosquito bites.

Although most of the interviewees knew that malaria was transmitted by mosquitoes, their understanding of the malaria vectors was not clear. Besides reporting bites from mosquitoes at night, the highest biting rates reported in the early evening were in Ubiyau and Sawanawa. From dusk until nightfall, many mosquitoes were collected in these villages, but most belonged to other genera rather than *Anopheles* (internal data not shown). In most households, mosquitoes were perceived as a problem both indoors and outdoors. Most of the interviewees mentioned that sleeping under mosquito nets was an alternative way to prevent mosquito bites. Other methods included spraying insecticides on house walls, making fire or smoke, using personal repellents, trimming bushes, and removing stagnant water around the houses. Since IRS was not fully and/or continuously implemented in all villages (Table S1-1D), the understanding of the IRS was also limited. In Ubiyau, which had the most IRS conducted, there was a better understanding of its usefulness. Meanwhile, ITNs and their function were better understood across all villages.

### Gaps in protection

The household survey highlighted several gaps in protection against *Anopheles* bites (Table 6). None of the surveyed homes had door or window screens, which would have helped limit mosquito entry. Screen use did not line up with socioeconomic status, but it did vary by village. Pitewi stood out, with about half of its households reporting screened doors and windows. Eaves were another weak spot being potential mosquito entry points. Sixty-four percent of homes had open eaves without screens, and only 17% had them partially screened. About one-third of households did not use any indoor mosquito repellent. This issue varied by site and by socioeconomic status, but it was still relatively small compared to the larger structural gaps.

**Table 6.**
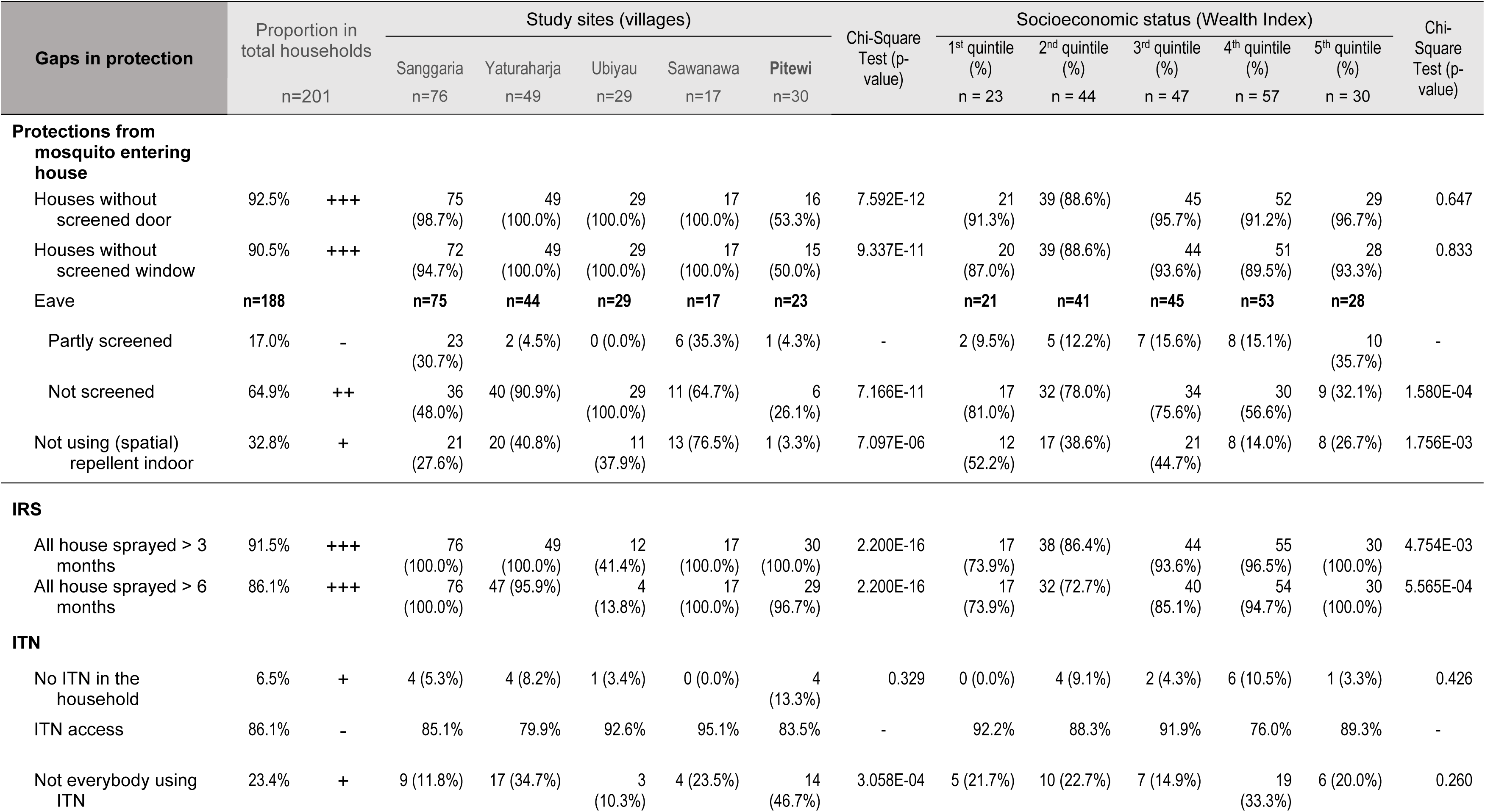
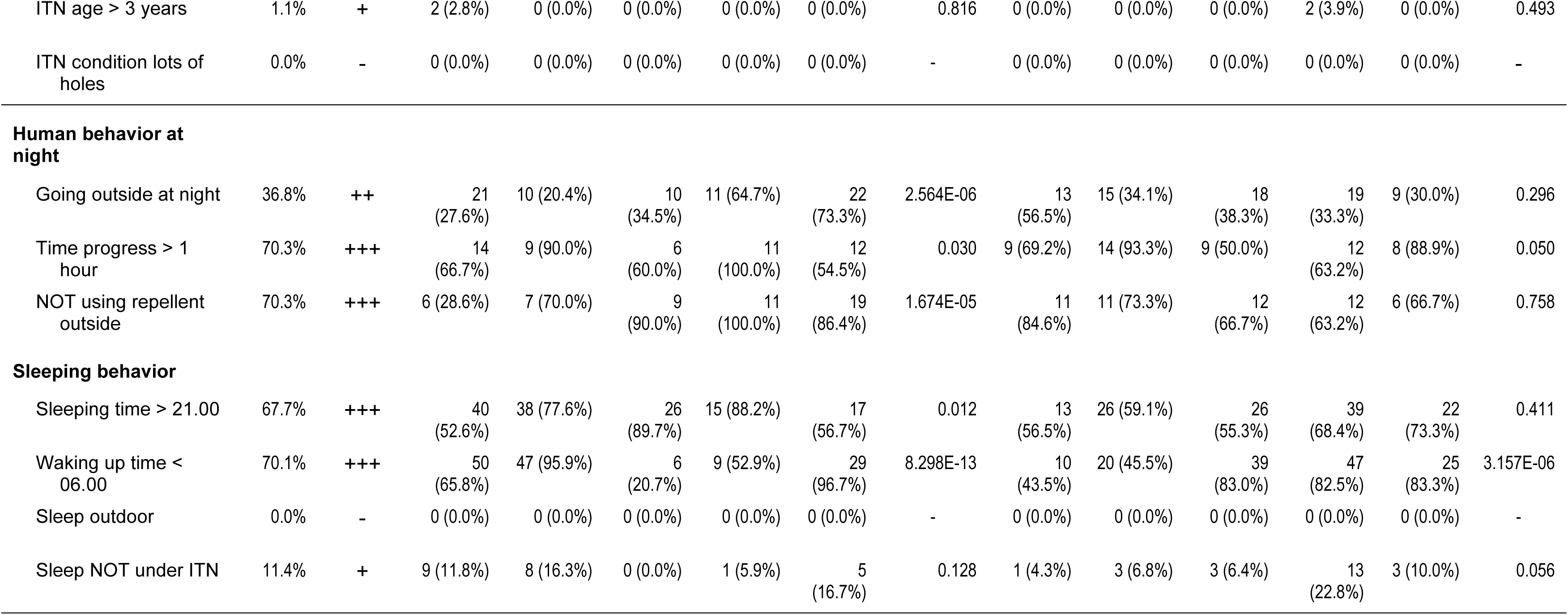
The list of gaps in protection identified in this study and their correlations with the study sites (villages) and the socioeconomic status of the households.

IRS coverage was a problem almost everywhere. Ubiyau was the exception, where 86.2% of homes had been sprayed at least once in the previous six months (Table S1-1D). ITN coverage, on the other hand, was high across all villages. The remaining gaps in protection were small and mostly practical issues, like nets older than three years or nets with multiple holes. Despite high ITN coverage, the proportion of all household members not using ITNs was quite high. Nighttime outdoor activity was another clear gap in protection. We categorized this as a moderate issue because many people went outside at night, stayed out for more than an hour, and rarely used any personal repellent during that time. This pattern was strongly linked to survey location but did not correlate with socioeconomic status. Sleeping patterns also created exposure windows. Most respondents went to bed after 21:00 and woke before 06:00, which overlapped with peak mosquito activity. These habits were associated with the study sites, and only wake-up time was related to socioeconomic status. Other factors such as sleeping outdoors and not sleeping under ITN were only minor issues during this study.

## DISCUSIONS

Malaria incidence in Keerom Regency has remained consistently high over the past decade (Fig 1), despite intensified malaria control efforts centered on three main strategies: early diagnosis and prompt treatment, provision of ITNs, and indoor residual spraying (IRS). Understanding the drivers of this persistent transmission requires not only epidemiological surveillance but also targeted entomological assessments. Previous rapid entomological surveys conducted across eight regencies in Papua, including Keerom, revealed ongoing malaria transmission both indoors and outdoors [7]. The present study, carried out in five villages of Keerom Regency, further confirms this dual transmission pattern, as reflected by the high entomological inoculation rate (EIR). Collectively, these findings underscore the value of rapid entomological assessments as practical tools for identifying key transmission dynamics in Papua [7,8], which is critical for tailoring vector control strategies effectively.

Molecular identification of *Anopheles* species is fundamental for characterizing the vectors responsible for malaria transmission, understanding their breeding habitats, and clarifying host-seeking behaviors. In line with previous observations [7,8], this study identified the Punctulatus group as the dominant vectors in Keerom, displaying both indoor and outdoor biting behavior with nearly equal frequency. Their primary habitats are located within and around residential areas. This evidence highlights a critical limitation of current control measures: interventions such as ITNs and IRS, which target indoor biting, are insufficient to interrupt transmission occurring outdoors. Mitigating outdoor malaria transmission requires coordinated efforts between local communities and technical personnel, particularly entomologists. Such efforts should include behavioral interventions to reduce outdoor exposure without protection during peak biting hours, community-driven larval source management, and entomologically guided interventions to target productive habitats.

Analysis of blood meals from mosquitoes collected during indoor resting revealed that members of the Punctulatus group, specifically *An. koliensis* and *An. punctulatus*, feed on both humans and animals (dogs). This finding demonstrates that while these species have a high human blood index (HBI), they also exhibit opportunistic feeding behavior. Similar observations have been reported in neighboring Papua New Guinea, where Punctulatus group species also opportunistically feed on animals [39–41]. This behavior could potentially be leveraged in vector control strategies, such as the use of animal-baited interventions to reduce human-vector contact [40,42,43]. However, multiple blood feeding on humans remains a concern, as it increases human-mosquito-human contact and can accelerate parasite development within the mosquito, ultimately enhancing overall malaria transmission potential [44,45].

Consistent with prior studies, the Punctulatus group was the predominant *Anopheles* taxa collected across all surveyed villages. Specifically, *An. koliensis*, *An. punctulatus*, and *An. hinesorum* (a member of the Farauti Complex) were the major species in lowland inland areas. *An. hinesorum* is largely associated with inland lowland river valleys and is the most widespread member of the Farauti Complex [46]. Although there are no reports confirming its role as a malaria vector in Papua, Indonesia, in Papua New Guinea it has been classified as a minor vector [47,48]. *An. bancroftii* is similarly considered a minor vector in Papua New Guinea [48], but its vectorial role in Papua, Indonesia, remains unproven. These findings highlight the need for localized entomological characterization to inform targeted interventions.

Human behavioral observations identified substantial gaps in protection against mosquito bites both indoors and outdoors (Table 6). Indoors, exposure primarily occurred before bedtime, between 18:00 and 21:00, exacerbated by the absence of mosquito screens on doors, windows, and eaves. Notably, only half of the population reported sleeping under ITNs. Outdoor exposure was largely associated with evening social activities, family gatherings, hunting, and garden attendance [29,49–51]. Larval habitat surveys indicated that ponds, ditches/gutters, and puddles/rain pools surrounding residential areas were key *Anopheles* breeding sites, with ponds being the most productive habitat type. Within these habitats, the presence of vegetation was significantly associated with higher larval densities.

The study identifies several actionable targets for malaria control in Keerom Regency. First, indoor interventions such as ITNs and IRS must be complemented with strategies addressing outdoor transmission, including community-led larval source management and behavior change to reduce nighttime outdoor exposure. Second, the opportunistic feeding behavior of the Punctulatus group suggests potential for animal-baited interventions to divert bites from humans. Third, improving house infrastructure, such as installing mosquito screens on doors, windows, and eaves, can reduce early evening indoor exposure. Finally, ongoing entomological monitoring should guide the spatial and temporal targeting of interventions to maximize impact.

Overall, the combination of entomological and human behavioral data highlights that malaria transmission in Keerom is sustained by both indoor and outdoor vector activity and is exacerbated by gaps in protection. Effective control in this setting will therefore require integrated strategies that combine conventional interventions targeting indoor transmission with measures addressing outdoor exposure and larval source management, informed by ongoing entomological monitoring.

## Limitations of the study

This study was limited by the small number of villages sampled, which may not capture the full geographic and ecological variability of Keerom Regency. The short duration of sampling may have missed seasonal fluctuations in Anopheles species and transmission dynamics. Mosquito collections and human behavioral observations were also limited in scope, potentially underestimating vector diversity, outdoor biting, and exposure patterns. Despite these constraints, the findings provide important insights for targeting malaria control interventions.

## CONCLUSIONS

Malaria transmission in Keerom Regency is sustained by both indoor and outdoor Anopheles activity, with the Punctulatus group as the dominant vectors. Gaps in human protection, particularly during early evening hours and outdoor activities, contribute to ongoing transmission. Effective control will require integrated strategies that combine conventional indoor interventions with measures targeting outdoor exposure, community-led larval source management, and behavior change. Opportunistic feeding on animals by key vectors also presents potential for novel interventions, while continued entomological monitoring is essential to guide and optimize control efforts.

## Data Availability

All data produced in the present work are contained in the manuscript

## ACKNOWLEDGEMENTS

We would like to express our gratitude to the residents of Sanggaria, Yaturaharja, Ubiyau, Sawanawa and Pitewi villages of Keerom Regency, Papua, those who participated directly or indirectly in this study, the local malaria cadres and public health center staffs who guided and assisted us in the fields. Special gratitude is also extended to the Keerom Regency Health Department.

We extend sincere appreciation to the National Research and Innovation Agency (BRIN) for research fundings in this study, i.e. Second Expedition and/or Exploration Funding program in 2022 (SK DFRI-BRIN No. 2860/II.7/HK.01.00/8/2022), and Rumah Program fundings from Research Organization of Health, BRIN (SK ORK-BRIN No. 23/III.9/HK/2023, No. 6/III.9/HK/2024, and No. 6/III.0/HK/2025) and Research Organization of Life Sciences and Environment, BRIN (SK ORHL-BRIN No. 1/III.5/HK/2024). Furthermore, we thank to the Eijkman Research Center for Molecular Biology, BRIN and Hasanuddin University for their continued support. This study is part of a doctoral program at Hasanuddin University, supported by the Degree by Research Program, BRIN.

